# Tripterygium glycosides as a potential treatment for CAR-T induced cytokine release syndrome: implication of monocyte depletion

**DOI:** 10.1101/2020.11.22.20232801

**Authors:** Zuqiong Xu, Fang Tian, Biqing Chen, Xiangtu Kong, Xingbin Dai, Jiang Cao, Pengjun Jiang, Jianxin Tan, Lu Lu, Xiachang Wang, Qi Lv, Di Kang, Miao Xu, Yingying Hu, Aiping Yang, Qian Wang, Zhong-Fa Yang, Xuemei Sun, Leyuan Ma, Lihong Hu, Xuejun Zhu

**Author notes:** Correspondence to: Dr. Xuejun Zhu (leading corresponding author), Department of Hematology, Jiangsu Province Hospital of Chinese Medicine/ the Affiliated Hospital of Nanjing University of Chinese Medicine, 155 Hanzhong Road, Qinhuai District, Nanjing 210029, China,; Dr. Lihong Hu, Jiangsu Key Laboratory for Functional Substance of Chinese Medicine, School of Pharmacy, Nanjing University of Chinese Medicine, Nanjing 210023, China,; Dr. Ma at David H. Koch Institute for Integrative Cancer Research, Massachusetts Institute of Technology Cambridge, Massachusetts 02139, United States,. Zuqiong Xu, Fang Tian, and Biqing Chen contributed equally to this article.

## Abstract

**Background:** Cytokine release syndrome (CRS) is a potentially life-threatening complication of chimeric antigen receptor T (CAR-T) cell therapy. Recent studies indicated critical roles of macrophages and monocytes in CAR-T induced CRS. Here, we report rapid dissipation of CAR-T induced CRS in two patients after receiving Tripterygium glycosides (TG). Effects of triptolide, the major active component of TG, on macrophages and monocytes were examined in animal models.

**Methods:** Two patients with CRS after CAR-T cell therapy (for hematological malignancy) received TG (50 mg, p.o.). Flow cytometry analysis and single cell RNA sequencing (scRNAseq) were conducted to examine the effects of TG on immune cells. Potential effects of triptolide were also examined *ex vivo* using patient-derived monocytes, as well as in mice.

**Findings:** Rapid alleviation of fever and cytokine storm was observed within 72 hours after TG treatment. Blood concentration of triptolide ranged from 21 to 154 ng/mL during treatment. Flow cytometry and scRNAseq showed selective depletion of monocytes with minimal impact on CAR-T cells in both patients. In *ex vivo* experiments with patient-derived monocytes, triptolide dramatically inhibited the synthesis of pro-inflammatory cytokines (e.g., IL-6, IL-10, and IP-10). Triptolide also rapidly and selectively depleted peritoneal concanavalin A activated macrophages and monocytes in mice.

**Interpretation:** TG could be a promising treatment for CAR-T induced CRS, as well as other diseases with similar mechanisms, e.g., hemophagocytic lymphohistiocytosis and COVID-19. Our preliminary findings require further verification with properly designed clinical trials.

## Introduction

Cytokine release syndrome (CRS), an acute systemic inflammatory response characterized by fever and multiple organ damages, is a life-threatening complication of chimeric antigen receptor T (CAR-T) cell therapy.^1–3^ Treatment for CRS include tocilizumab and corticosteroids.^4^ However, only a proportion of patients with CRS respond to tocilizumab, and corticosteroids could ablate the CAR-T cell persistence and efficacy.^4,5^ Recent studies indicate that macrophages and monocytes are the predominant source of cytokines that mediate CAR-T induced CRS.^6,7^ Selective depletion of monocytes thus represent a promising approach to manage CAR-T induced CRS.

Tripterygium glycosides (TG), a chloroform/methanol extract from *Tripterygium wilfordii* Hook F (LeiGongTeng or Thunder God Vine), are effective in the treatment of a variety of autoimmune and inflammatory diseases.^8^ Triptolide, the major active component of TG, has been shown to induce apoptosis in monocytes,^9^ and inhibit macrophage activation via TGF-Beta Activated Kinase 1 Binding Protein 1.^10^

We recently used TG in a patient with hemophagocytic lymphohistiocytosis (HLH), and achieved rapid response (data not published). Considering the overlapping pathological features of HLH and CAR-T induced CRS, we use TG in two patients who developed CRS after CAR-T cell therapy. Rapid alleviation of CRS was observed in both patients. Flow cytometry analysis and single cell RNA sequencing (scRNAseq) demonstrated selective depletion of monocytes in these patients. We then conducted a series of *ex vivo* experiments with patient’s blood and animal experiments, and showed that triptolide could decrease the synthesis of pro-inflammatory cytokines by depleting macrophages and monocytes.

## Methods

### TG in the two patients with CAR-T induced CRS

Patient 1was enrolled in a trial of anti-CD19-CAR-T cell therapy in patients with recurrent/refractory CD19^+^ leukemia or lymphoma (Chinese Clinical Trial Registry number ChiCTR-OIC-17013081). Patient 2 was enrolled in a trial of anti-CD19/BCMA-CAR-T cell therapy in patients with relapsed/refractory multiple myeloma (ChiCTR1900028098).

Humanized anti-BCMA CAR and anti-CD19 CAR lentiviral vectors were constructed as previously described (Fig. 1).^11,12^ T lymphocytes were isolated using anti-CD3 immuno-magnetic beads. CAR-T cells were prepared as previously described.^13^ At 2 days prior to CAR-T infusion, the two patients received fludarabine (at a dose of 30 mg/m^2^ of bodysurface area) and cyclophosphamide (at a dose of 600 mg/m^2^) to deplete lymphocytes. Patient 1 received 1.31×10^6^ CD3^+^ cells/kg (0.52×10^6^ CD19-specific CAR-T cells/kg) in a single dose. Patient 2 received 0.75×10^6^ CD3^+^ cells/kg (0.25×10^6^ CD19-specific CAR-T cells/kg), followed by another infusion of 4.69×10^6^ CD3^+^ cells/kg (1.72×10^6^ BCMA-specific CAR-T cells/kg). Clinical effects for CAR-T cell therapy were evaluated as described in the trial protocol (ChiCTR-OIC-17013081 and ChiCTR1900028098). The cytokine release syndrome evaluation criteria proposed by Neelapu and colleagues was adopted.^14^

**Figure 1.**
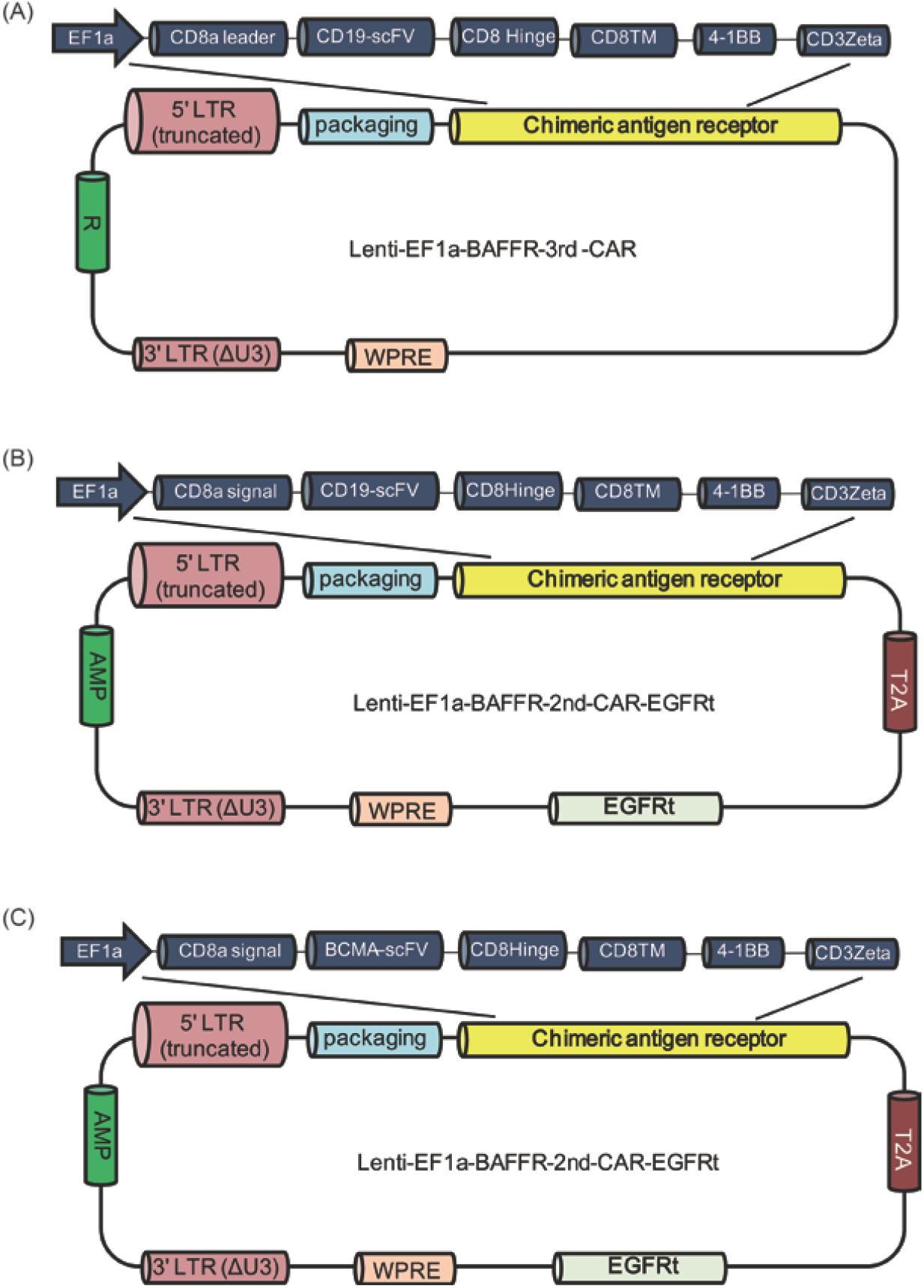
Design of CAR-T lentiviral vectors. The structure of anti-CD19-CAR for Patient 1 is shown in panel A. Shown in panel B and C are the structures of anti-CD19-CAR (B) and anti-BCMA-CAR (C) for Patient 2, which include a truncated EGFR (EGFRt) segment.

TG was given as tablets (Zhejiang DND Pharmaceutical Co.,Ltd, Zhejiang, China; 10 mg per tablet) at a dose of 10 mg three times a day (TID) with a total dose at 50 mg.

### qPCR analysis

Genomic DNA was isolated from peripheral blood mononuclear cells (PBMCs). To quantify the copy numbers per unit DNA, a 5-point standard curve was generated by spiking 10^2^ to 10^6^ copies of CTL019 lentiviral plasmid into 100 ng of non-transduced control genomic DNA. qPCR analysis was performed using SYBR-Green to detect the integrated CD19 CAR transgene sequence using 100 ng genomic DNA per time-point. Primers specific for a non-transcribed genomic region upstream of the CDKN1A and WPRE were:

#### CDKN1A

forward: gaaagctgactgcccctatttg

reverse: gagaggaagtgctgggaacaat

#### WPRE

forward: ccttttacgctatgtggatacg

reverse: ccaggatttatacaaggaggaga

### Soluble cytokines analysis

Serum concentration of 12 representative soluble cytokines was determined using a multiplexed-microsphere-based flow cytometry immunoassay (Raisecare Biological Technology Co., Ltd., Shandong, China). Data were acquired on a Navios Flow Cytometer (Beckman Coulter, Inc., CA, USA) and analyzed using LEGENDplex v8.

### Minimal residual disease analysis

Minimal residual disease (MRD) was monitored 24 (Patient 1) or 18 (Patient 2) days after CAR-T cell infusion using a Navios Flow Cytometer and analyzed with Kaluza (Beckman Coulter). The antibody panel for detection of MRD in Patient 1 included anti-CD45-PerCP/Cy5.5 (clone: H100), anti-CD19-PC7 (clone: 89B), anti-CD34-APC (clone: 581), anti-CD13-PE (clone: SJ1D1), anti-CD33-PE (clone: D3HL60.251), anti-HLA-DR-FITC (clone: Immu-357), anti-CD10-PerCP/Cy5.5 (clone: HI10a), anti-CD20-BV421 (clone: 2H7). The panel in Patient 2 included anti-CD45-PC5.5 (clone: J.33), anti-CD19-PC7 (clone: 89B), anti-CD38-ECD (clone: LS198-4-3), anti-c-kappa-FITC & anti-c-lambda-PE (polyclonal), anti-CD27-APC (clone: 1A4CD27), anti-CD138-BV421 (clone: MI15), anti-CD56-BV510 (clone: NCAM16.2).

### Triptolide concentration in blood

An ultra-performance liquid-chromatography tandem mass spectrometry (UPLC-MS/MS) method was developed and optimized to determine triptolide concentration in blood.^15–18^ The reference standard (triptolide purity ≥ 98%) was provided by Sigma-Aldrich (St. Louis, USA). The blood samples was extracted with ethyl acetate.^16^ Hydrocortisone (purity ≥ 98%; Shanghai Ruiyong Biotechnology Co., Ltd., Shanghai, China) was usedas an internal standard (IS).^15,17^ The UPLC-MS/MS system consisted of an UPLC system (UPLC 20A, Shimadzu Corporation, Kyoto, Japan) and a triple quadrupole tandem mass spectrometry (QTRAP™ 5500 MS system, AB Sciex Corporation, CA, USA) coupled with an electrospray ion source. Selectivity and specificity of the method, and precision and accuracy of intra-day and inter-day analysis were evaluated and were acceptable. The detection was repeated five times.

### Effects of triptolide in patient-derived cells

This experiment was carried out in Patient 2 from one clinical trial (ChiCTR1900028098) and five patients from another trial (ChiCTROIC-17011272) that intended to evaluate the safety and efficacy of anti-BCMA CAR-T and anti-CD19 CAR-T therapy for relapsed and refractory multiple myeloma. The experiments were conducted in triplicate for the sample from each patient.

Heparinized peripheral blood was mixed with complete medium at equal volume, and incubated with triptolide (final concentration: 0, 1, 3, 10, 30 or 100 ng/mL) for 24 hours prior to multi-parameter flow cytometry immunophenotyping. Approximately 1×10^6^ total cells per condition were stained in 100 μL PBS for 30 minutes on ice using antibodies at concentrations recommended by manufacturer.

The antibodies in the multi-parameter immunophenotyping included: anti-CD3-ECD (clone: UCHT1), anti-CD14-AF700 (clone: RMO52), anti-HLA-DR-PE (clone: B8.12.2). Flow cytometry was performed on a Navios Flow Cytometer with data analyzed by Kaluza. Gating for monocytes in patient’s blood was live cells (FSC/SSC) > CD14^+^ vs. HLA-DR^+^. For *ex vivo* culture, gating was live cells (FSC/SSC) > SSC low events > CD3^+^ for T cells, and live cells (FSC/SSC) > SSC low events > CD14^+^ for monocytes.

### mRNA sequencing

PBMCs were treated with triptolide (30 ng/mL) or vehicle for 12 hours. Total RNA was extracted and sequenced on NovaSeq6000 (Illumina, CA, USA). Raw data were preprocessed by FASTQ, aligned to *Homo sapiens* reference genome (HISAT2 index: Ensembl GRCh38 genome_tran) by HISAT2 v.2.0.5, and analyzed with StringTie v.1.3.3b and edgeR.

### Single cell RNA sequencing

PBMCs were treated with triptolide (10 or 30 ng/mL) or vehicle for 12 hours. Single cells were separated and scRNA-seq libraries were generated for the 3’ scRNA-seq on a 10X genomics platform with v3 chemistry and sequenced on a NovaSeq 6000. Sequencing data were aligned to human genome GRCh38 and cells were counted by Cellranger (v.3.1.0). Seurat v.3 was used for quality control, treatment integration, and data visualization. The filter criteria were: genes expressed in at least three cells; cells with expression of no less than 200 genes; cells with < 30% mitochondria genes and < 30% HBB-related genes. T-SNE was used to reduce dimensions. Bulk gene expression was calculated by averaging single cell data in each cell subgroups to mimic bulk-RNA sequencing. Heatmaps and violin plots were generated by R 4.0.1. CD14^+^ cells were defined as CD14 expression ⩾ 2 reads; T cells were defined as CD3D expression ⩾ 2 reads; CD16^+^ cells were defined as FCGR3A expression ⩾ 2 reads.

### Effect of triptolide on concanavalin A-activated mouse peritoneal monocytes/macrophages

Male SPF-grade C57BL/6 mice (6-8 weeks of age) were challenged with concanavalin A (ConA) (C5275, Sigma) at 4mg/kg (i.p.) to activate peritoneal monocytes/macrophages. Starting from the next day, mice randomly received triptolide (200 or 400 ng/kg, i.p.) or vehicle for 3 consecutive days (n=5). A group of healthy mice receiving neither ConA nor triptolide was included as an additional control. At 24 hours after the last treatment, mice were sacrificed, abdominal cavity was washed with 5mL PBS containing 2 mM EDTA, and cells were collected by centrifugation. Flow cytometry was conducted using fluorescent labeled antibodies (F4/80-FITC and Ly6c-Pe-Vio770).

## Results

### TG rapidly mitigated CAR-T induced CRS

Patient 1 received surgical resection of primary ovarian cancer followed by six courses of adjuvant chemotherapy. She was then diagnosed with acute B lymphoblastic leukemia and achieved complete remission after treatments four years later. Leukemia relapsed 15 months later, with no response to chemotherapy (e.g., vindesine, epirubicin, and cyclophosphamide). Seven days after CAR-T cell infusion, she developed recurrent fever with a body temperature of up to 40.0°C (Fig. 2A) but not responded to acetaminophen, accompanied by hypoxia (Table 1), hypotension (Fig. 2B), and elevated serum C-reactive protein (CRP), procalcitonin (PCT), lactate dehydrogenase (LDH) (Fig.2C), and multiple inflammatory cytokines (Fig.2D). Her CRS grade was 2. At two days after CRS emergence, she received 10 mg TG TID for a total dosage at 50 mg. Her body temperature returned to normal (Fig. 2A) and most cytokines (e.g., IL-6, IL-10, IFN-γ) dramatically decreased (Fig. 2D) after 40 hours, accompanied by gradual decrease in PCT, LDH, and CRP levels (Fig. 2C). Her liver and renal function remained normal throughout the TG treatment (Fig. 2E). Remission with minimal residual disease (MRD) and hematopoietic recovery was achieved on the 24th day (Fig. 3A). Her leukemia relapsed after nine months.

**Table 1.**
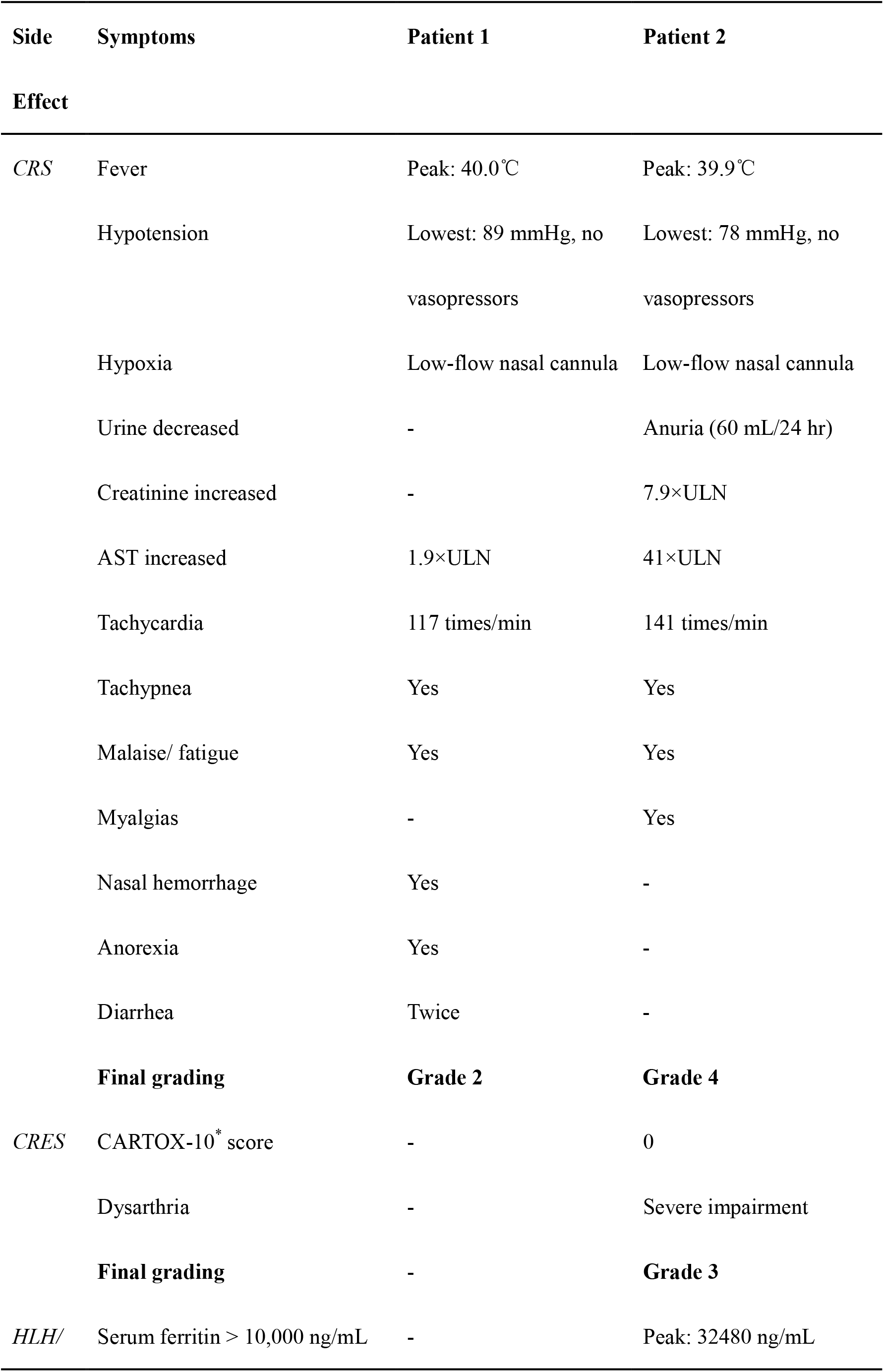
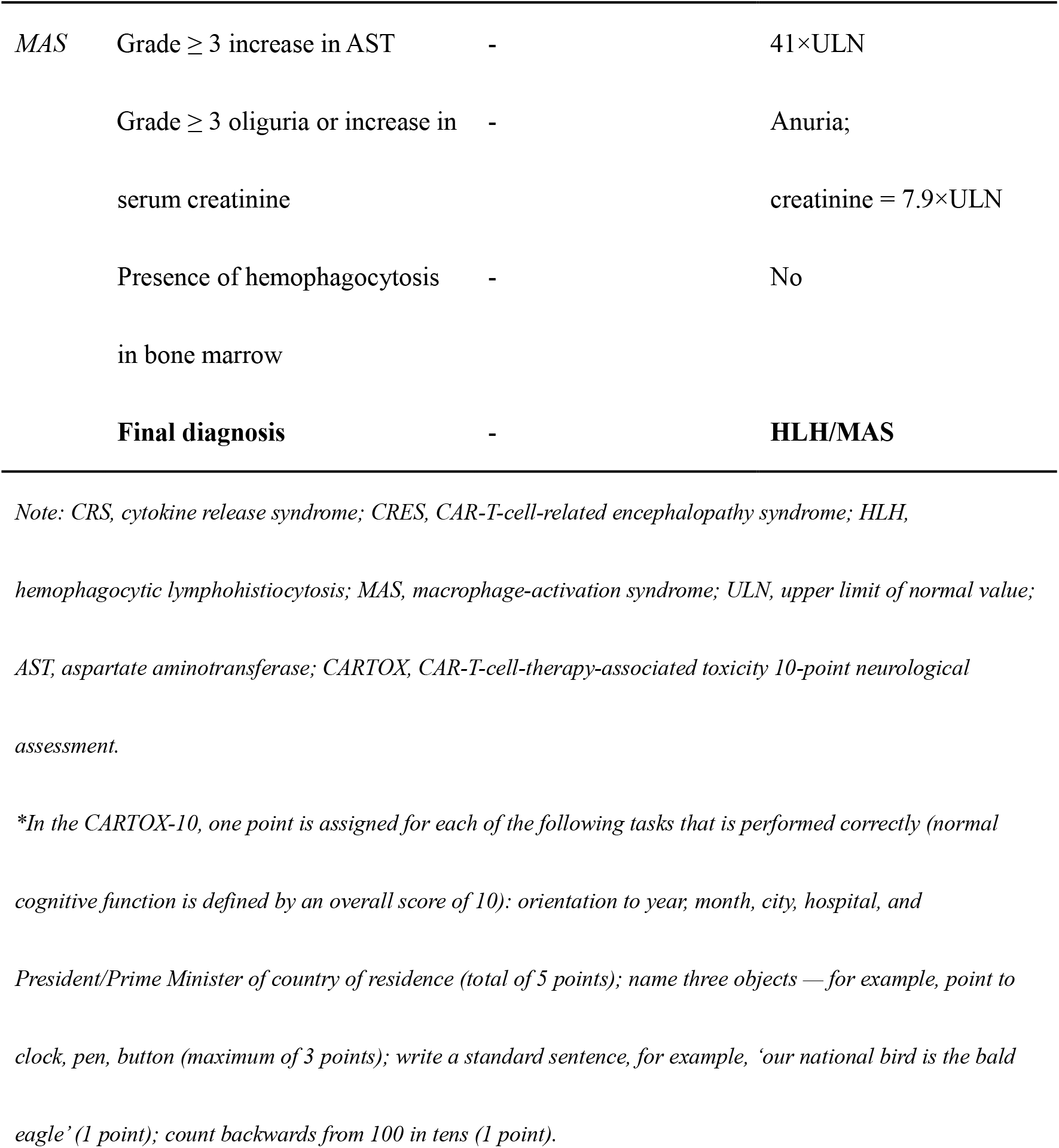
Assessment of CAR-T therapy related side effects

**Figure 2.**
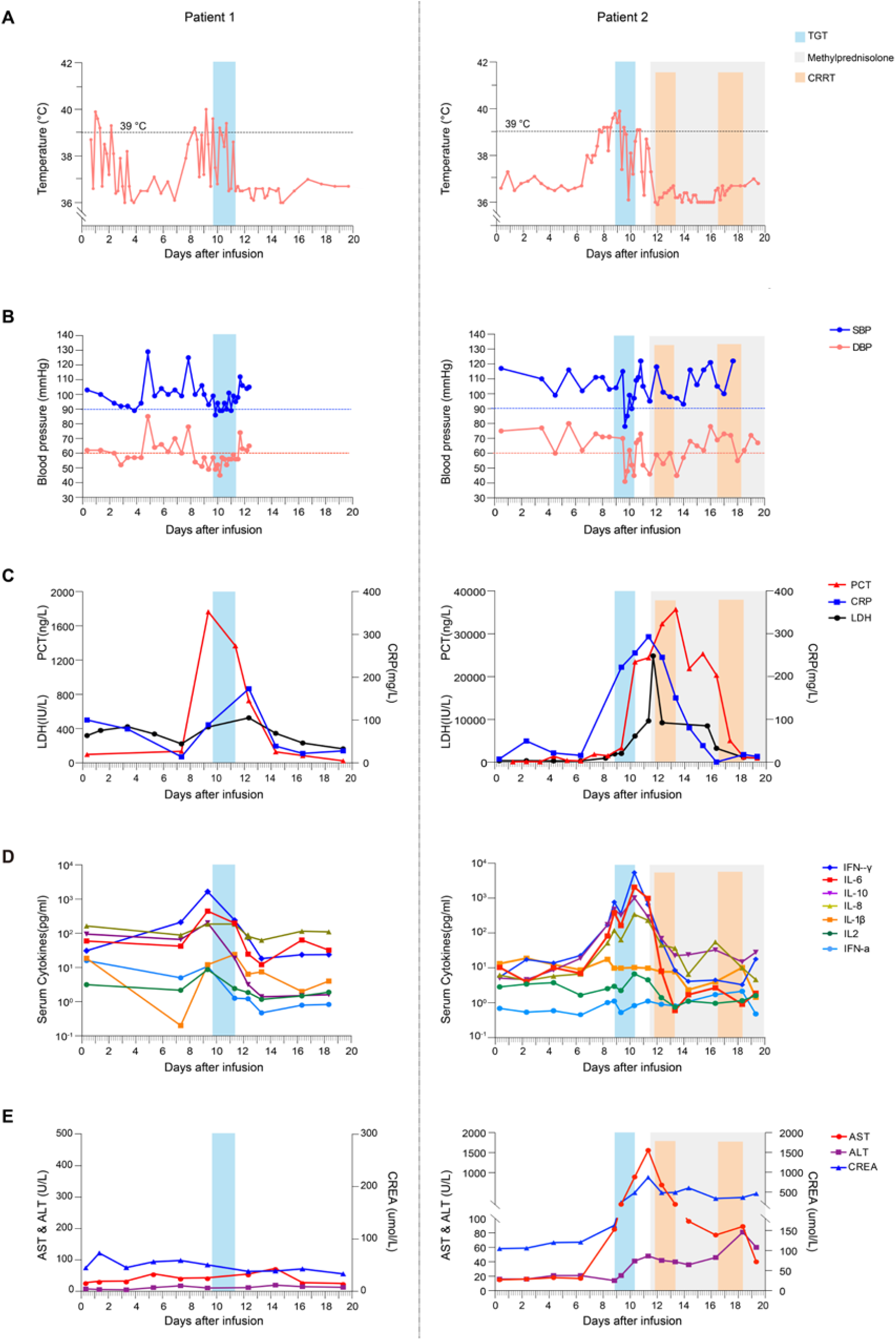
Clinical responses to TG treatment in two patients with CAR-T induced CRS. Day one: first CAR-T cell infusion. A: body temperature. B: blood pressure. DBP, diastolic blood pressure; SBP, systolic blood pressure. Hypotension is defined as systolic blood pressure < 90 mmHg. C: procalcitonin (PCT), lactate dehydrogenase (LDH), and C-reactive protein (CRP). D: serum cytokine levels measured by flow cytometry. E: liver and kidney function, as reflected by serum alanine aminotransferase (ALT), aspartate aminotransferase (AST), and serum creatinine (CREA). TG treatment, continuous renal replacement therapy (CRRT), and methylprednisolone treatment are indicated by colored blocks.

**Figure 3.**
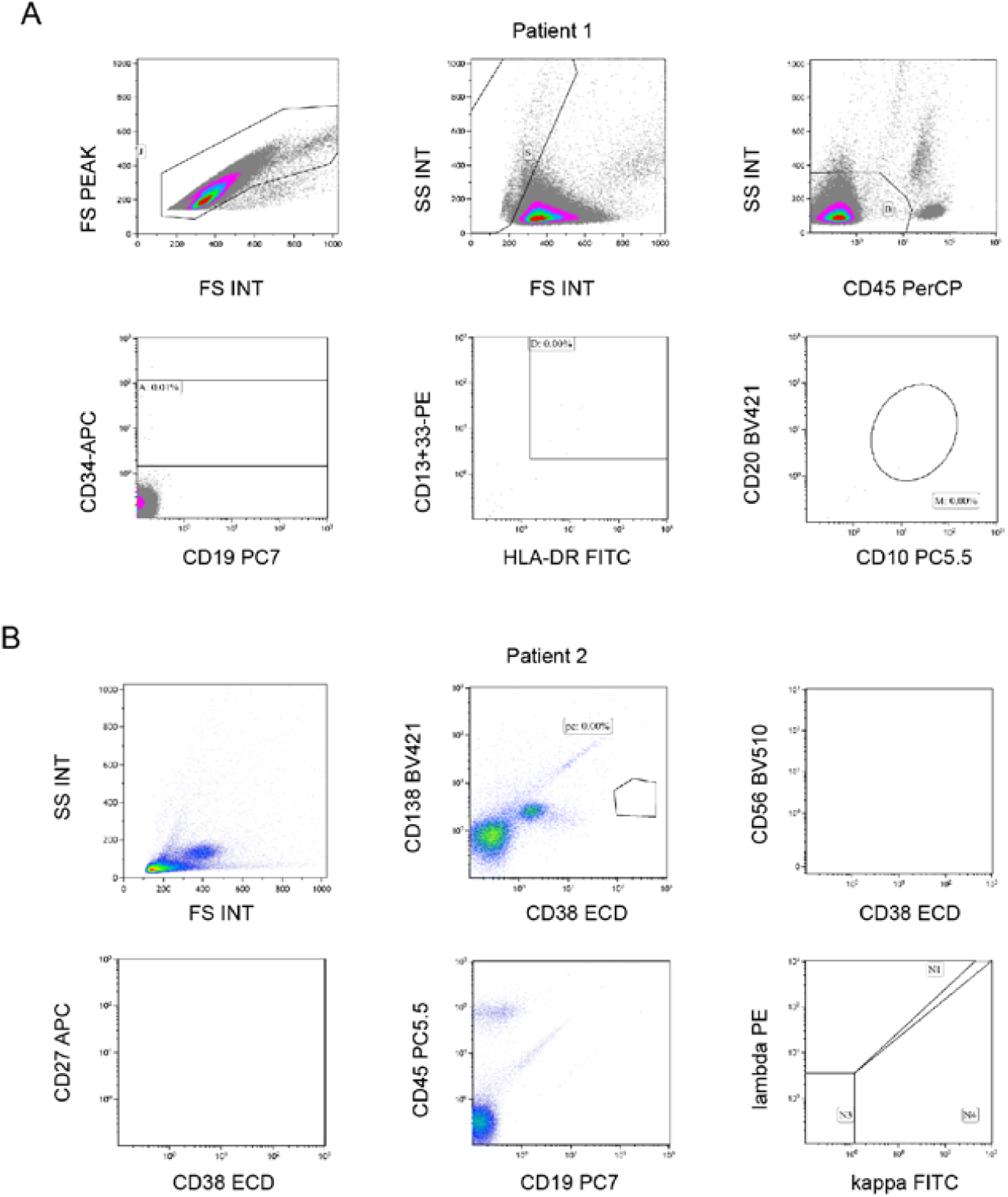
Minimal residual disease (MRD) after CAR-T therapy. A: Patient 1. The number of B-ALL cells expressing CD19, CD34, CD10, CD20, CD13, CD33, HLA-DR, and CD45 on the 24th day after CAR-T therapy was less than 10^−4^, confirming MRD negativity. B: Patient 2. The number of multiple myeloma cells expressing CD138, CD38, CD56, CD27, CD45, CD19, c-kappa and c-lambda on the 18th day after CAR-T therapy was less than 10^−4^, confirming MRD negativity.

Patient 2 with multiple myeloma (IgD-lambda) achieved a partial response to lenalidomide, bortezomib, and dexamethasone (RVD) two years before presentation. The baseline karyotype was 46, XY, +add(1)(p11), -3, del(3)(q13), add(9)(q34), del(11)(p12), add(14)(q32), -17, 18, add(20)(q11), add(22)(q11), +2mar, inc[20]. Complex karyotype and interphase fluorescence *in situ* hybridization showed cytogenetic abnormalities in 55% multiple myeloma cells and 1q21 amplification, suggesting poor prognosis. The disease progressed six months later. He received three cycles of RVD with cyclophosphamide, followed by treatment with lenalidomide, pomalidomide, melphalan, vindesine, epirubicin, and arsenious acid, but bone destruction and anemia exacerbated. A bone marrow aspiration showed 52% of total nucleated cells were abnormal plasma cells. Six days after CAR-T cell infusion, he developed persistent fever with a body temperature of up to 39.9°C (Fig. 2A) but no response to acetaminophen, accompanied by hypoxia (Table 1), elevated CRP, PCT, LDH (Fig. 2C), and multiple inflammatory cytokines (Fig. 2D). Diagnoses of grade 4 CRS, grade 3 CAR-T cell-related encephalopathy syndrome (CRES) and HLH were established (Table 1). Starting from two days after CRS emergence, he received 10 mg TG TID at 50-mg total dosage. His persistent fever relieved after 20 hours (Fig. 2A). Majority of serum cytokines, e.g., IL-6, IL-10, IFN-γ, decreased significantly after TG treatment before receiving methylprednisolone. Alanine aminotransferase (ALT), aspartate aminotransferase (AST), and creatinine (CREA) remained elevated. He was transferred to the intensive care unit (ICU) on the 12th day after CAR-T infusion, and received methylprednisolone and continuous renal replacement therapy (CRRT) treatment (Fig. 2E). After seven days of treatment, CRS and CRES returned to normal. Remission with MRD and hematopoietic recovery was achieved on the 18th day (Fig. 3B). He died of multidrug-resistant *Klebsiella pneumonia* sepsis on the 27th day (Table 2 and 3).

**Table 2.** Antibiotic susceptibility of the bacteria isolated from blood samples of Patient 2

| Antibiotics name | MIC (µg/mL) | Sensitivity |
| --- | --- | --- |
| Ampicillin/Sulbactam | ≥ 32.0 | R |
| Piperacillin/Tazobactam | ≥ 128.0 | R |
| Ceftriaxone | ≥ 64.0 | R |
| Imipenem | ≥ 16.0 | R |
| Gentamicin | ≥ 16.0 | R |
| Ciprofloxacin | ≥ 4.0 | R |
| Compound sulfamethoxazole | ≤ 20.0 | S |
| Cefotetan | ≥ 64.0 | R |
| Ceftazidime | ≥ 64.0 | R |
| Cefepime | ≥ 64.0 | R |
| Aztreonam | ≥ 64.0 | R |
| Amikacin | ≥ 64.0 | R |
| Levofloxacin | ≥ 8.0 | R |
| Tobramycin | ≥ 16.0 | R |

*Note: R, resistant; S, sensitive; MIC, Minimum inhibitory concentration. Sample was collected at 24 days after*
*CAR-T cell infusion and 14 days after the last TGT administration. Test results were reported after Patient 2's*
*death. The bacteria identified from blood samples of Patient 2 was **Klebsiella pneumoniae subsp. pneumoniae**.*

**Table 3.**
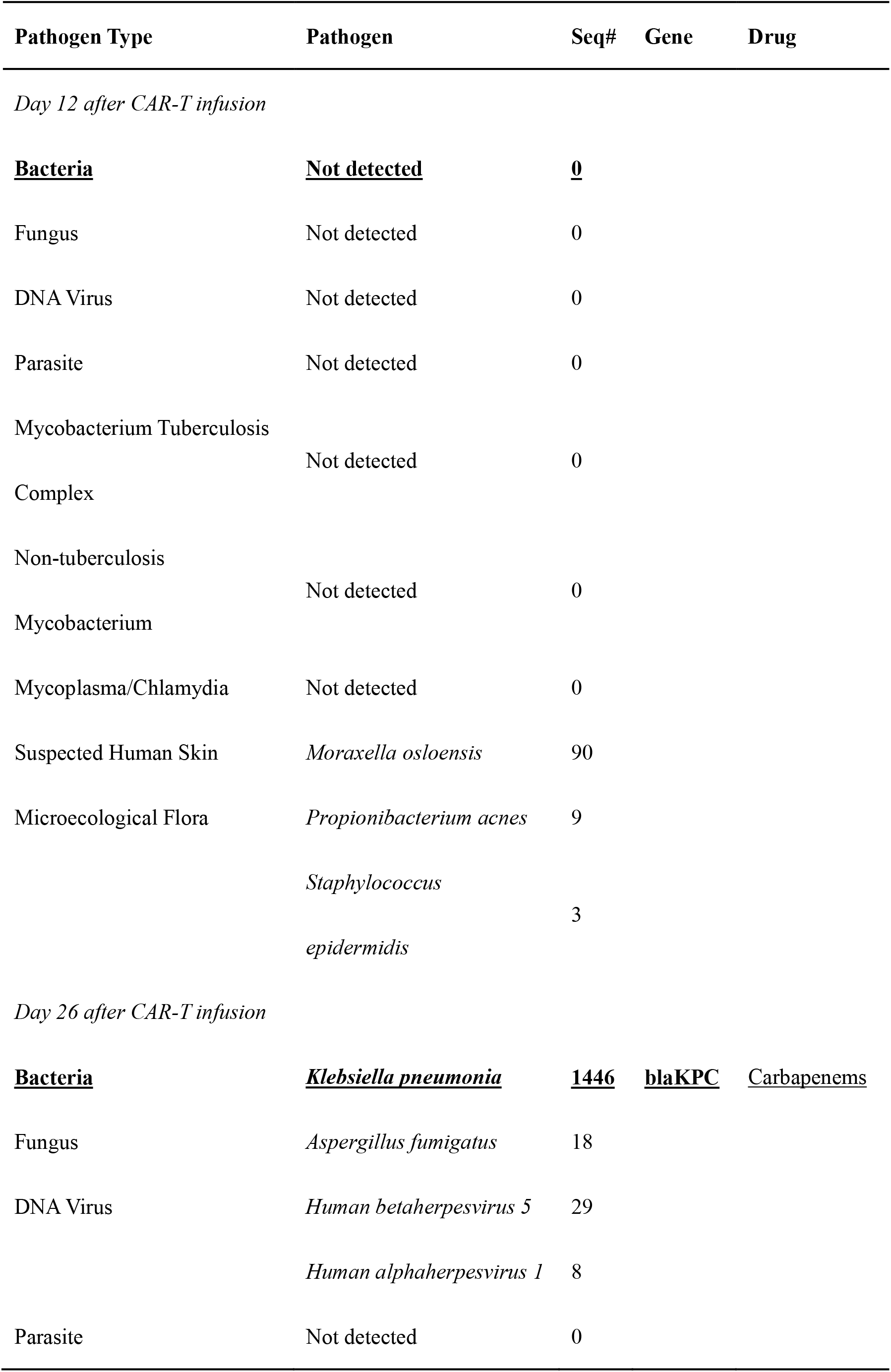
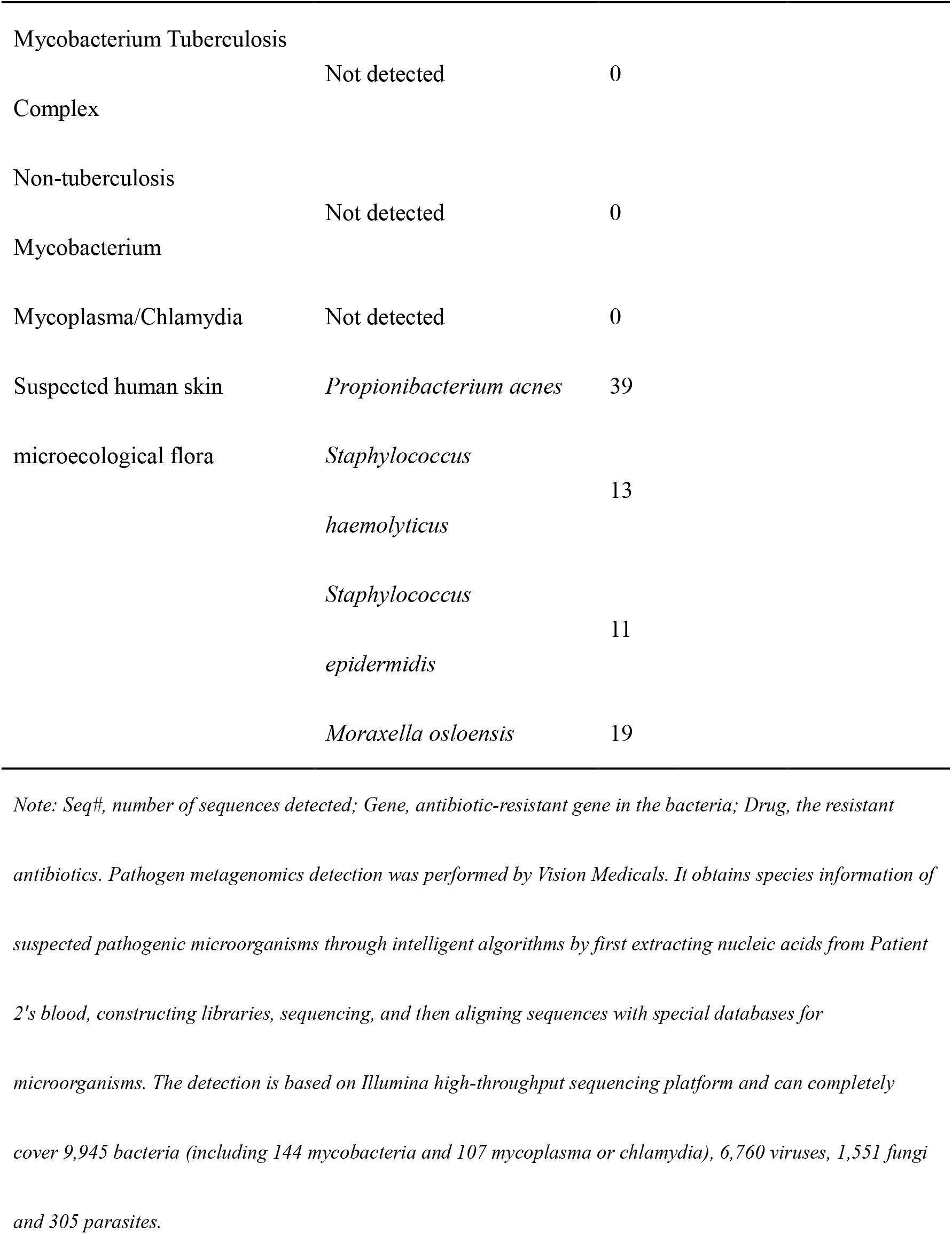
Pathogens metagenomics analysis for Patient 2

As ingredients vary among TG tablets from different pharmaceutical manufacturers, we focused on the one used in our hospital (Zhejiang DND Pharmaceutical Co.,Ltd). A separate study in patients with autoimmune diseases (mainly rheumatoid arthritis) confirmed minimal liver and renal toxicity for this brand (7.69% grade 1 renal impairment after TG treatment, Fig. 4), consistent with a previous meta-analysis showing low toxicity.^19^

**Figure 4.**
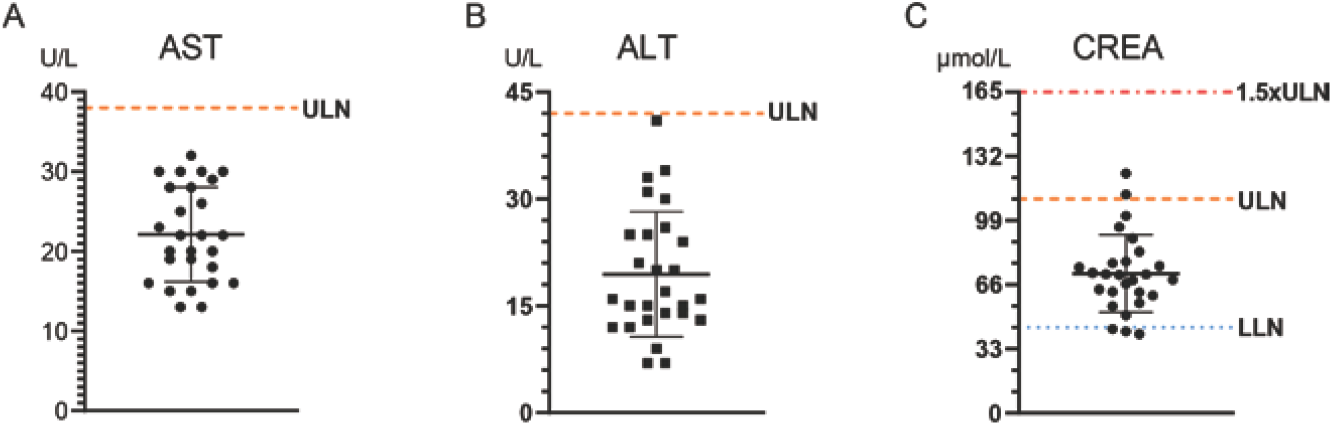
Effect of TG treatment on liver and renal functions in patients with autoimmune diseases. As ingredients vary among TG tablets from different pharmaceutical manufacturers, we focused on the one used in our hospital (Zhejiang DND Pharmaceutical Co.,Ltd). 26 patients who received TG for a variety of autoimmune diseases for at least one week were included in the assessment: 19 with rheumatoid arthritis, 2 with Sjogren’s syndrome, 3 with chronic glomerulonephritis,1with ankylosing spondylitis, and 1with gouty arthritis. Dosage included 10 mg TID (n=5), 20 mg BID (n=13), 20 mg TID (n=6), 30 mg BID (n=1), and 40 mg BID (n=1). A: serum aspartate aminotransferase (AST). B: alanine aminotransferase (ALT). C: creatinine (CREA). The upper limit of normal value (ULN) is indicated by orange line, the 1.5×ULN is indicated by red line, and the lower limit of normal value (LLN) is indicated by blue line. One patient (an elder) has taken TG for years and had renal dysfunction prior to hospitalization (CREA = 125.5 μmol/L); her CREA decreased somewhat after receiving TG during hospitalization. Only one patient with normal measures before hospitalization had elevated CREA (112.4 μmol/L) after receiving TG during hospitalization.

### Plasma triptolide concentration in patients

Triptolide is a major active component of TG, and often used for pharmacokinetic studies of TG.^20^ We developed a UPLC-MS/MS method to determine plasma triptolide concentration in patients. The sensitivity and accuracy are shown in Fig. 5. As an initial optimization, we analyzed blood samples collected from six patients treated with TG for autoimmune or inflammatory diseases. Plasma triptolide concentration in these patients was 57.4 ∼ 149.9 ng/mL (Fig. 5E). Plasma triptolide after the first oral dosing was 154.4 ng/mL and 34.5 ng/mL in Patient 1 and Patient 2, respectively (Fig. 5E), and undetectable three days after the last dosing.

**Figure 5.**
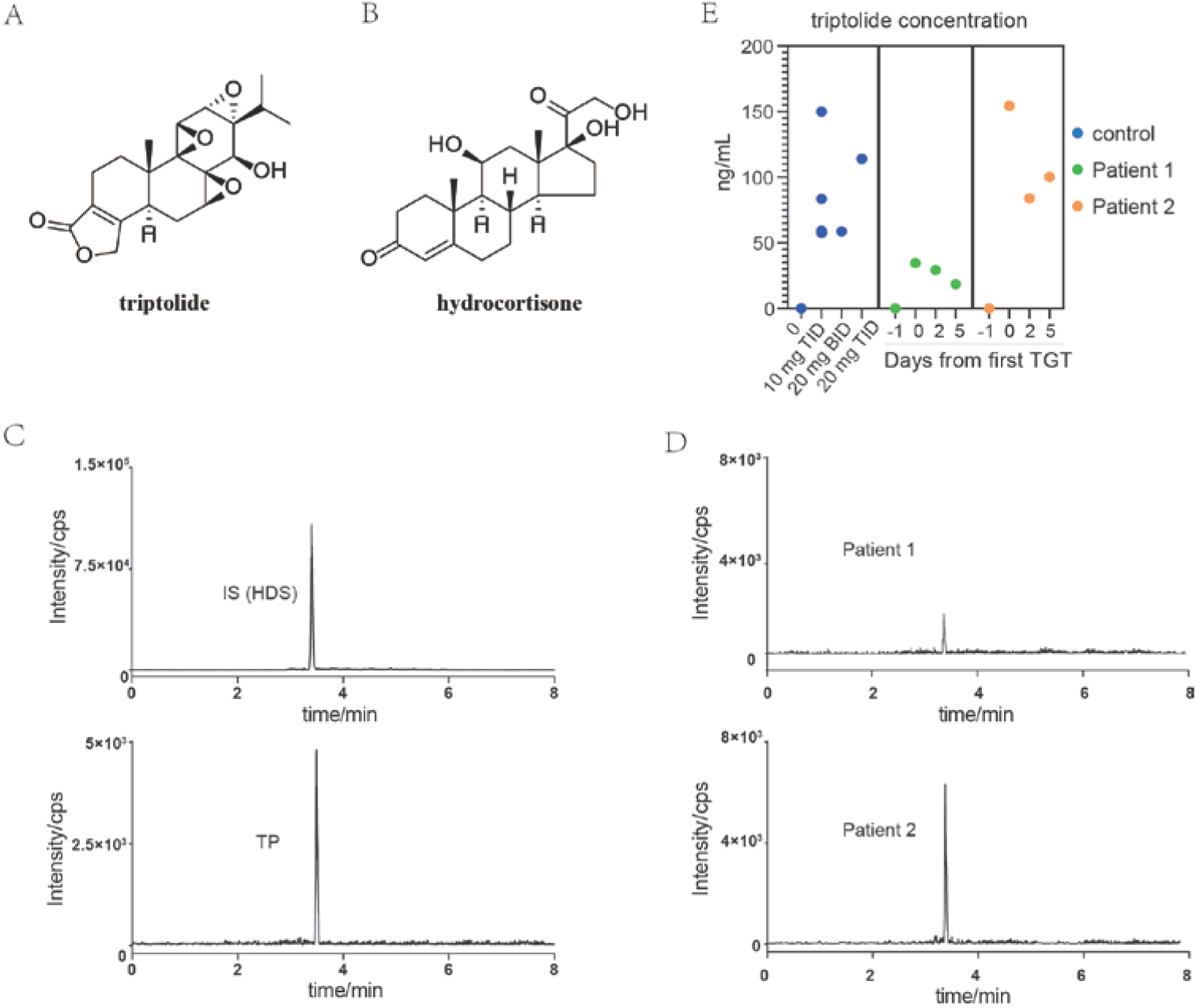
HPLC-MS/MS analysis of triptolide in human plasma. A: the structure of triptolide (TP). B: the structure of the internal standard (IS) - hydrocortisone (HDS). C: representative UPLC-MS/MS chromatogram of blank plasma spiked with 100 ng/mL HDS and 100 ng/mL TP. D: patients plasma samples collected at two hours after receiving the first TG dosing. E: triptolide in human plasma during TG treatment. Each dot represents one sample. The dark blue dots represent patients with inflammatory autoimmune diseases. Six patients received TG at three doses: 10 mg TID, 20 mg BID, and 20 mg TID. Three additional patients with rheumatoid arthritis who did not receive TG are included as negative control. The green and orange dots are samples from Patient 1 and Patient 2, one day before TG treatment (Day -1), 2 hours after receiving the first TG dosing (Day 0), 2 hours before receiving the last TG tablet (Day 2), and three days after the last treatment (Day 5).

### TG selectively depleted monocytes without affecting CAR-T Cells in patients

The number of monocytes (CD14^+^ HLA-DR^+^ cells) in the peripheral blood rapidly decreased after TG treatment (Fig. 6A and 6B). Quantitative real time-PCR showed no reduction in the number of CAR-T cells after TG treatment in both patients (Fig. 6C). There was no MRD in either patient, suggesting minimal influence on CAR-T cells (Fig. 3).

**Figure 6.**
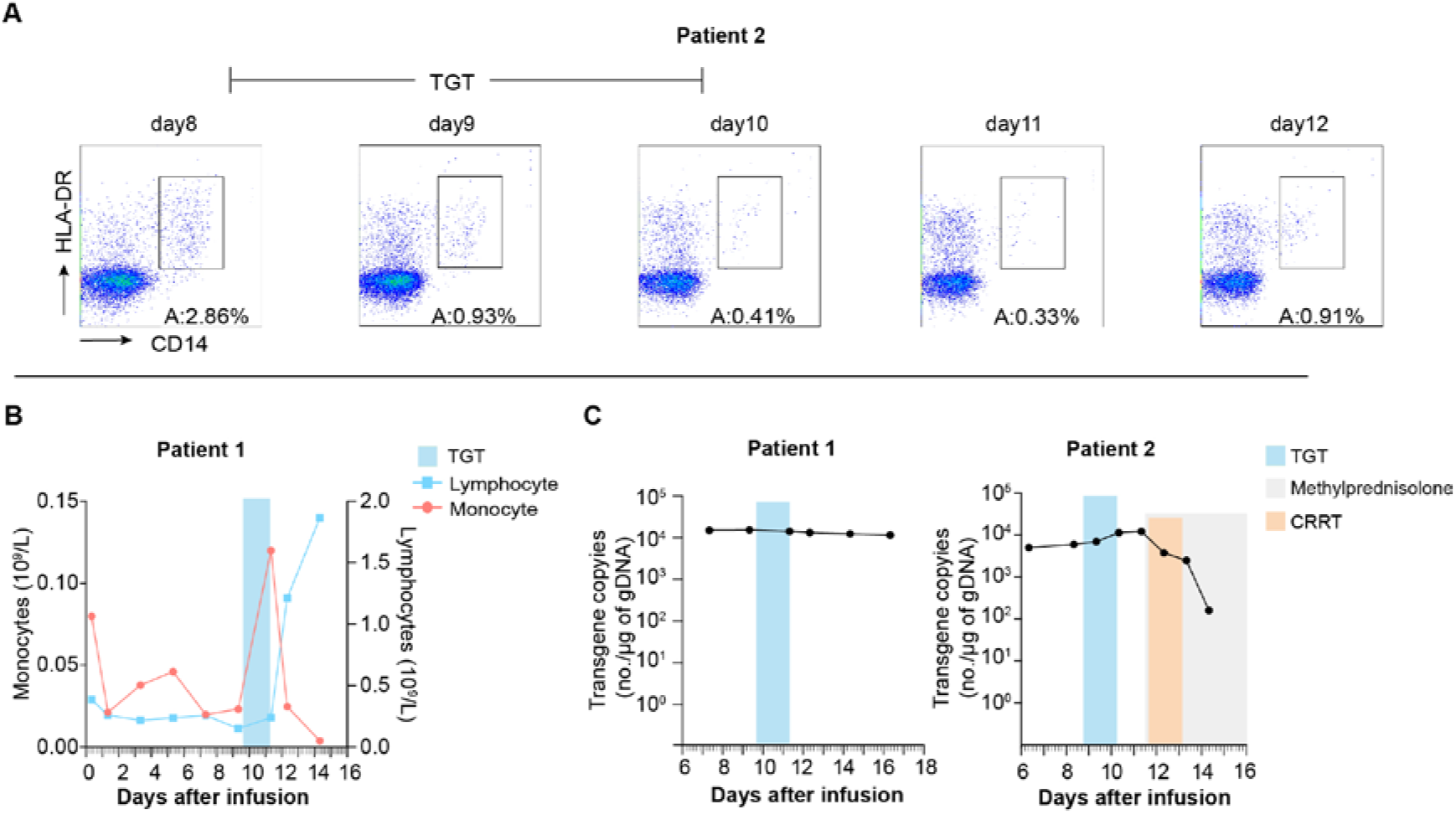
Effect of TG treatment on monocytes, lymphocytes and CAR-T cells in patients. A: peripheral blood samples were immunophenotyped by flow cytometry to determine the percentages of monocytes (CD14^+^ HLA-DR^+^ cells) in white blood cells for Patient 2. B: absolute number of monocytes and lymphocytes in Patient 1. Monocytes in the peripheral blood decreased after TG treatment in both patients. In Patient 1, monocytes in the peripheral blood dropped to near zero three days after the last TG treatment. C: copy numbers (no.) of transgenic DNA per μg of genomic DNA (gDNA), as determined with quantitative real-time PCR. The copies of transgenic DNA remained consistent during TG treatment. TG treatment, continuous renal replacement therapy (CRRT), and methylprednisolone treatment are indicated by colored blocks.

### *Ex vivo* triptolide treatment selectively depleted monocytes in patient’sblood

Flow cytometry analysis indicated selective depletion of monocytes in a dose-dependent manner with negligible effect on T cells when peripheral blood collected from the patients were treated with triptolide *ex vivo* (Fig. 7). At 100 ng/mL, triptolide removed the majority of monocytes.

**Figure 7.**
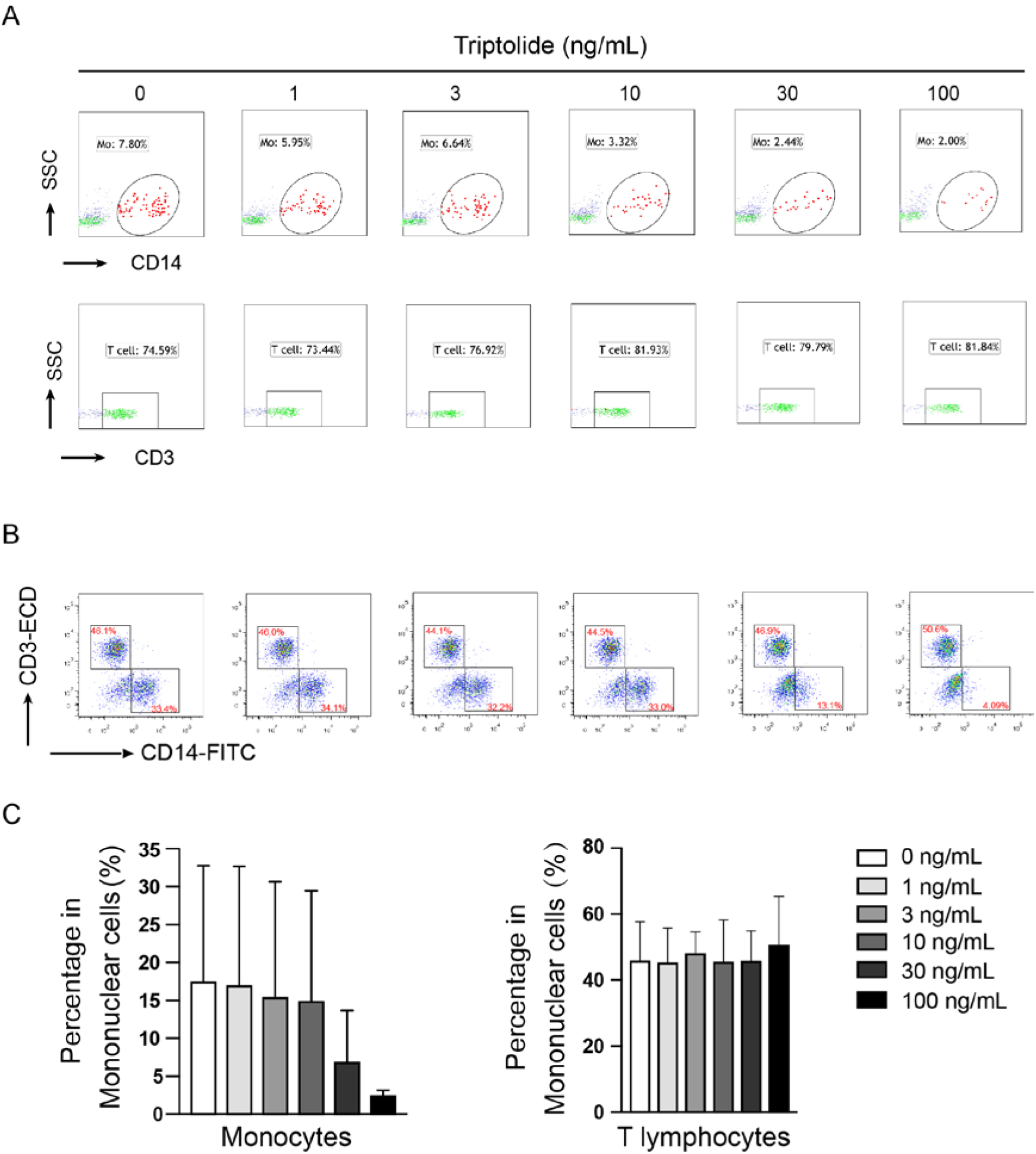
Effect of triptolide on the quantity of monocytes and T cells *in vitro*. On the eighth day after CAR-T cell infusion (immediately prior to TG treatment), PBMCs were collected from Patient 2. PBMCs from patients were treated with triptolide (1, 3, 10, 30, or 100 ng/mL) for 24 hours prior to flow cytometry analysis for monocytes (CD14^+^ cells) and total T cells (CD3^+^ cells). Representative flow cytometry result showing selective and dose-dependent decrease in monocytes for Patient 2 (A) and five patients with CRS (B). (C) Statistical summary based on five patients with CRS.

### *Ex vivo* triptolide treatment decreased cytokine synthesis in patient’s monocytes

*Ex vivo* triptolide treatment decreased the gene expression of most cytokines known to be involved in CRS (Fig. 8A). Nine out of the 17 CRS-related cytokines decreased substantially after triptolide exposure, e.g., IL-10, IP-10, and IL-12p70. Pathways involved in cytokine signaling, translation regulation, and lymphocyte chemotaxis were significantly inhibited (Fig. 8B).

**Figure 8.**
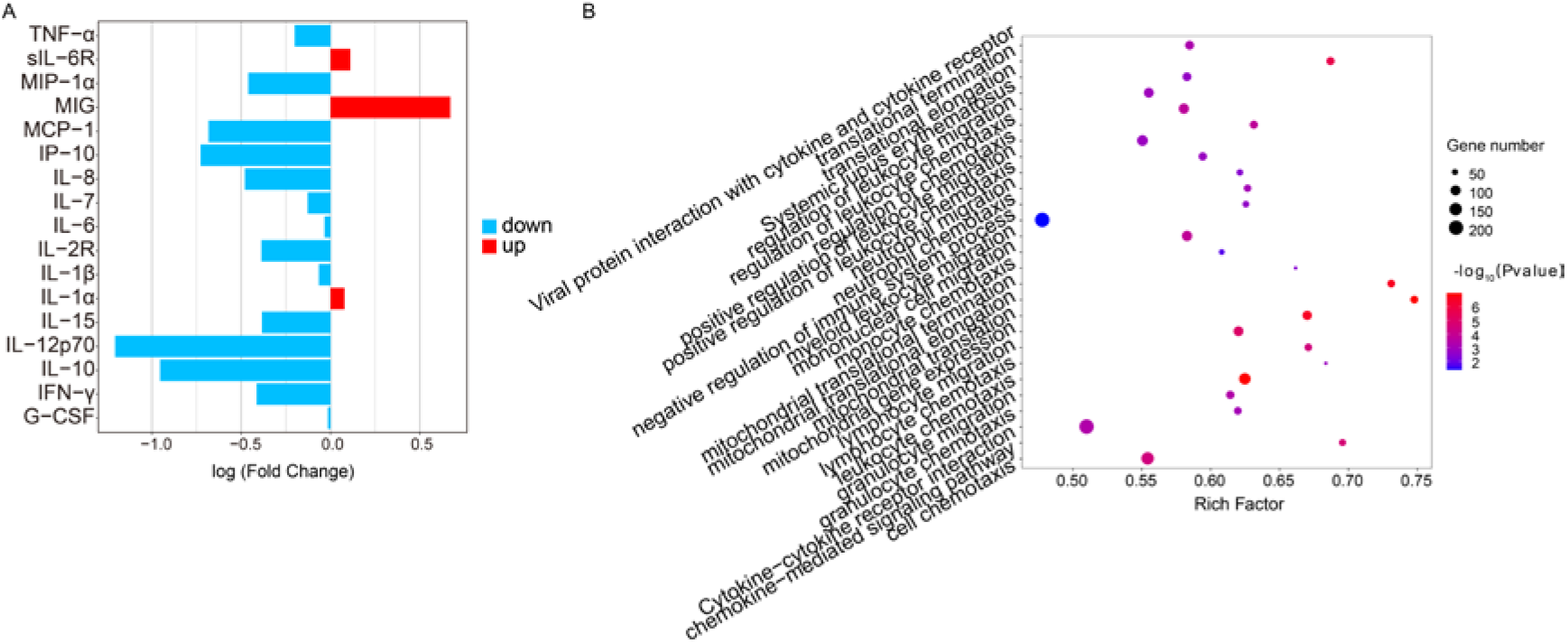
Effects of triptolide on mRNA expression in cultured monocytes from one CAR-T patient. (A) Gene expression of CRS-related cytokines. The bar represents the log10 transformed fold change in gene expression. Blue: down-regulation; red: up-regulation. (B) Significantly affected pathways revealed by gene enrichment analysis and transcript enrichment analysis.

### *Ex vivo* triptolide treatment selectively depleted CD14^+^CD16^+^ monocytes

Comprehensive profiling of PBMCs using scRNA-seq revealed that triptolide selectively depleted CD14^+^CD16^+^ monocytes without affecting CD14^+^CD16^-^ monocytes and CD3^+^ T cells (Fig. 9A and 9B). Notably, triptolide treatment dramatically decreased the expression of most pro-inflammatory cytokines in CD14^+^CD16^+^ monocytes (Fig. 9C), and particularly CRS-related cytokines, such as IL-6, IL-10, and IP-10 (Fig. 9D).

**Figure 9.**
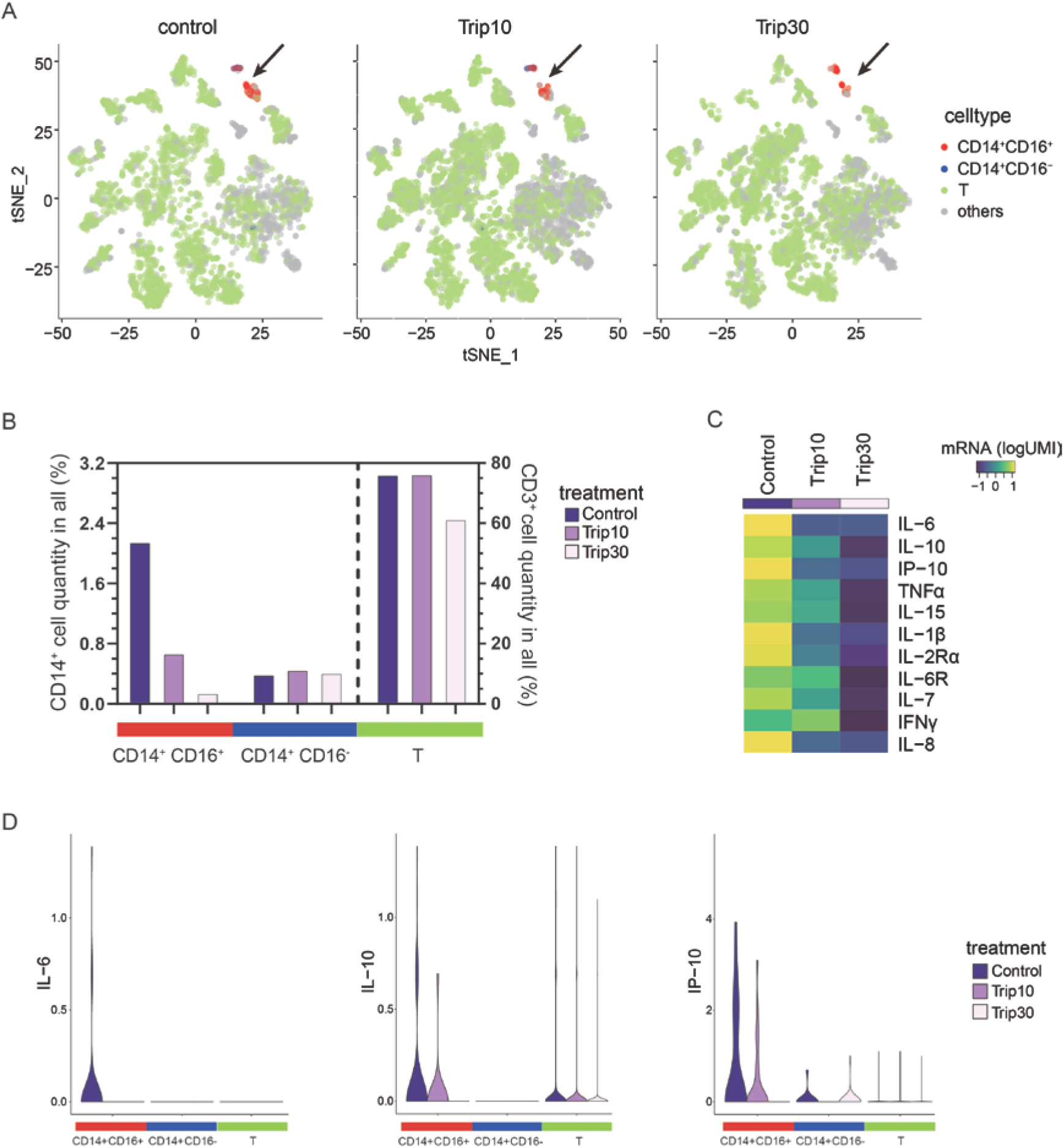
Single-cell RNA sequencing of triptolide-treated PMBCs. PBMCs collected from Patient 2 immediately prior to TG treatment were treated for 12 hours with triptolide (10 or 30 ng/mL) or vehicle for 12 hours prior to single-cell RNA sequencing. A: two subtypes of monocytes (CD14^+^ C16^+^, CD14^+^ CD16^-^) and T cells (CD3^+^) in t-SNE plots. Left: control; middle and right: triptolide at 10 and 30 ng/mL, respectively. Each dot represents one cell. B: the percentage of the two subtypes of monocytes and T cells in the whole sample. White: control; grey and black: triptolide at 10 and 30 ng/mL, respectively. C: the mean expression of genes encoding CRS-related cytokines in the whole blood sample. mRNA counts are determined as log10-transformed unique molecular identifiers (logUMI) and normalized across samples for each gene. D: selected cytokines that responded to triptolide treatment. The genes encoding IL-6, IL-10, and IP-10 were mainly expressed in CD14^+^ CD16^+^ monocytes: their expression decreased with increasing concentration of triptolide.

### *In vivo* triptolide treatment selectively depleted peritoneal monocytes/macrophages in ConA-treated mice

Triptolide (200 and 400 ng/kg) significantly reduced the number of peritoneal macrophages and monocytes in mice challenged with ConA (Fig. 10).

**Figure 10.**
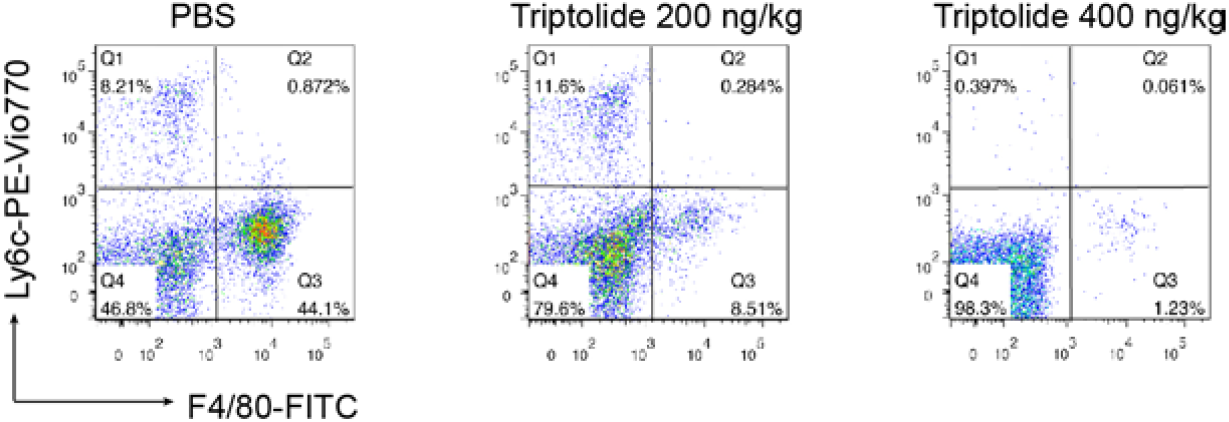
Effect of intra-peritoneal injection of triptolide on peritoneal monocytes/macrophages in mice challenged with concanavalin A (ConA). For each figure, monocytes are represented by Ly6c^high^ F4/80^-^ cells, macrophages are represented by Ly6c^low^ F4/80^+^ cells. Mice treated with ConA have increased number of macrophages, but when treated with triptolide, the number of monocytes/macrophages decreased in a dose-dependent manner.

## Discussion

CAR-T therapy represents an important milestone in cancer treatment,^2,3^ but is associated with potentially life-threatening CRS.^1,4,12^ Glucocorticoids are the first-choice for CRS but could expedite the clearance of CAR-T cells.^21,22^ Such a disadvantage is reflected by the rapid reduction of the proportion of CAR-T cells in peripheral blood after corticosteroid treatment in Patient 2 in the current study. MRD was not detected in this patient, but adverse impact on long-term remission could not be ruled out due to short follow-up. Besides, the cause of death in this patient (multi-drug resistant bacterial infection) could be associated with the use of corticosteroid. Tocilizumab is effective for CRS, but therapeutic efficacy varies across patients,^23^ likely due to the removal of only IL-6 but not other pro-inflammatory cytokines. There has been many recent development efforts, including modifying CAR-T to reduce the incidence of CRS,^24^ neutralizing and removing GM-CSF *in vivo*,^25^ and inhibiting CAR-T cells with dasatinib,^26^ but need to be verified by more large-scale clinical trials.

Studies in animal models suggested monocytes as a major source of cytokines in CRS. Removal of monocytes in mice with liposomal clodronate has been shown to decrease CRS incidence and mortality,^6^ but translation into human use is problematic since there is no drugs approved for human use that could selectively deplete monocytes. Etoposide is capable of removing activated monocytes and has been used to treat HLH, but with limited efficacy and, more importantly, the potential to inhibit hematopoiesis.^27^

TG is approved by the China Food and Drug Administration (Z32021007) as an anti-inflammatory and immunomodulatory agent,^8^ and to treat a variety of diseases, including rheumatoid arthritis, Crohn’s disease and ulcerative colitis.^28^ TG significantly reduces inflammatory cytokines (e.g., IL-6, IL-1, and TNF-α),^29^ but the mechanism remains unclear. In a previous study, IL-6 was primarily produced by monocytes and macrophages, and not CAR-T cells.^30^ In previous studies by this research group and others, triptolide has been shown to induce apoptosis in monocytes and monocyte-derived dendritic cells at concentration relevant to human use.^31,32^ A study by Titov et al. suggested that triptolide could target ERCC Excision Repair 3 and produce biological functions through inhibiting the transcription process by RNA polymerase.^33^ TAB1 has also been reported to be a target of triptolide: treatment of mouse macrophages with 30 nM triptolide inhibits the TAK1 kinase activity.^10^ In the current study, we showed that triptolide could selectively deplete peritoneal macrophages and monocytes in mice challenged with ConA. Considering the finding of selective depletion of monocytes in patients receiving TG, we speculate that the rapid abrogation of CRS in the two patients in the current study is due to depletion of monocytes/macrophages although the human study part was not designed to investigate the potential cause-effect relationship.

In conclusion, the 2 patients receiving TG for CAR-T induced CRS experienced rapid dissipation of CRS. Such a response was associated with significantly decreased monocytes in peripheral blood. *In vitro* experiments using cells from the patients and *in vivo* experiments using ConA-treated mice confirmed selective depletion of macrophages and monocytes, with negligible effect on CAR-T cells. These findings are clearly preliminary and need to be verified by further studies in animal models and human patients with CRS induced by CAR-T therapy, and perhaps as well as CRS associated with other conditions, for example, HLH and COVID-19.^34,35^

## Data Availability

All data are available upon request by emailing to.

## Contributions

X Zhu and L Hu designed the whole study. The cases were initially seen by Z Xu. Z Xu, X Kong, X Dai, and X Zhu identified the cases and planned the clinical assessments. F Tian, B Chen, L Hu, and X Zhu planned the laboratory assessments. Z Xu, X Kong, X Dai, L Lu, Y Hu, Q Wang, and J Cao collected clinical data. Z Xu, X Kong, X Dai, P Jiang, M Xu, Y Hu, and X Zhu performed clinical assessments. B Chen, X Wang, Q Lv, D Kang, A Yang, and L Hu examined triptolide concentration in blood. F Tian did PCR. F Tian, B Chen, and X Zhu did mice experiments, *in vitro* culture and prepared cells for single cell sequencing. F Tian and B Chen analyzed the data. F Tian, B Chen, L Ma, and X Zhu interpreted the data. B Chen, J Tan, X Sun, X Zhu did the literature research. B Chen, J Tan, L Ma and X Zhu drafted the manuscript. F Tian, B Chen, J Tan, ZF Yang, L Ma, and X Zhu edited the manuscript.

## Acknowledgments

We thank Juhua Yu for sample collection and processing; Yixun Wu for flow cytometry analysis; Min Wu and Kang Wang for cytokines measurement. This study was supported in part by the Science and Technology Development Program of Jiangsu Province-Clinical Frontier Technology (grant Nos. BE2016809, BE2017639), the Nanjing Science and Technology Development Program (grant Nos. 201503011), the Priority Academic Program Development of Jiangsu Higher Education Institutions (PAPD) (grant Nos. 035062002003c), and the development program of Jiangsu Provincial Hospital of Chinese Medicine (grant Nos.Y19030, Y19066).

## References

1. Liu, D. & Zhao, J. Cytokine release syndrome: grading, modeling, and new therapy. J Hematol Oncol11, 121 (2018).

2. Grupp, S. A. et al. Chimeric antigen receptor-modified T cells for acute lymphoid leukemia. N. Engl. J. Med.368, 1509–1518 (2013).

3. Porter, D. L., Levine, B. L., Kalos, M., Bagg, A. & June, C. H. Chimeric Antigen Receptor–Modified T Cells in Chronic Lymphoid Leukemia. N. Engl. J. Med.365, 725–733 (2011).

4. Riegler, L. L., Jones, G. P. & Lee, D. W. Current approaches in the grading and management of cytokine release syndrome after chimeric antigen receptor T-cell therapy. Ther. Clin. Risk Manag.15, 323–335 (2019).

5. Teachey, D. T. et al. Identification of predictive biomarkers for cytokine release syndrome after chimeric antigen receptor T-cell therapy for acute lymphoblastic leukemia. Cancer Discov.6, 664–679 (2016).

6. Norelli, M. et al. Monocyte-derived IL-1 and IL-6 are differentially required for cytokine-release syndrome and neurotoxicity due to CAR T cells. Nat. Med.24, 739–748 (2018).

7. Giavridis, T. et al. CAR T cell-induced cytokine release syndrome is mediated by macrophages and abated by IL-1 blockade letter. Nat. Med.24, 731–738 (2018).

8. Han, R., Rostami-Yazdi, M., Gerdes, S. & Mrowietz, U. Triptolide in the treatment of psoriasis and other immune-mediated inflammatory diseases. Br. J. Clin. Pharmacol.74, 424–436 (2012).

9. Yuan, K. et al. Application and mechanisms of triptolide in the treatment of inflammatory diseases-a review. Front. Pharmacol.10, 1–12 (2019).

10. Lu, Y. et al. TAB1: A target of triptolide in macrophages. Chem. Biol.21, 246–256 (2014).

11. Garfall, A. L. et al. Chimeric antigen receptor T cells against CD19 for multiple myeloma. N. Engl. J. Med.373, 1040–1047 (2015).

12. Yan, Z. et al. A combination of humanised anti-CD19 and anti-BCMA CAR T cells in patients with relapsed or refractory multiple myeloma: a single-arm, phase 2 trial. Lancet Haematol.6, e521–e529 (2019).

13. Cao, J. et al. Potent anti-leukemia activities of humanized CD19-targeted Chimeric antigen receptor T (CAR-T) cells in patients with relapsed/refractory acute lymphoblastic leukemia. Ultrasound Obs. Gynecol.50, 776–780 (2006).

14. Neelapu, S. S. et al. Chimeric antigen receptor T-cell therapy-assessment and management of toxicities. Nat. Rev. Clin. Oncol.15, 47–62 (2018).

15. Zhang, Y. et al. Influence of verapamil on pharmacokinetics of triptolide in rats. Eur. J. Drug Metab. Pharmacokinet.41, 449–456 (2016).

16. Du, X. et al. Simultaneous determination of seven effective components of Tripterygium glycosides in human biological matrices by ultra performance liquid chromatography–triple quadrupole mass spectrometry. J. Chromatogr. B Anal. Technol. Biomed. Life Sci.1113, 1–13 (2019).

17. Gong, X., Chen, Y. & Wu, Y. Absorption and metabolism characteristics of triptolide as determined by a sensitive and Reliable LC-MS/MS Method. Molecules20, 8928–8940 (2015).

18. Shao, F. et al. Pharmacokinetic study of triptolide, a constituent of immunosuppressive Chinese herb medicine, in rats. Biol. Pharm. Bull.30, 702–707 (2007).

19. Ren, D., Zuo, C. & Xu, G. Clinical efficacy and safety of Tripterygium wilfordii Hook in the treatment of diabetic kidney disease stage IV: A meta-analysis of randomized controlled trials. Medicine (Baltimore).98, e14604 (2019).

20. Brudno, J. N. & Kochenderfer, J. N. Recent advances in CAR T-cell toxicity: Mechanisms, manifestations and management. Blood Rev.34, 45–55 (2019).

21. Davila, M. L. et al. Efficacy and toxicity management of 19-28z CAR T cell therapy in B cell acute lymphoblastic leukemia. Sci. Transl. Med.6, 224ra25 (2014).

22. Brentjens, R. et al. CD19-targeted T cells rapidly induce molecular remissions in adults with chemotherapy-refractory acute lymphoblastic leukemia. Sci Transl Med5, 177ra38 (2013).

23. Le, R. Q. et al. FDA Approval Summary: Tocilizumab for Treatment of Chimeric Antigen Receptor T Cell-Induced Severe or Life-Threatening Cytokine Release Syndrome. Oncologist23, 943–947 (2018).

24. Ying, Z. et al. A safe and potent anti-CD19 CAR T cell therapy. Nat. Med.25, 947–953 (2019).

25. Sterner, R. M. et al. GM-CSF inhibition reduces cytokine release syndrome and neuroinflammation but enhances CAR-T cell function in xenografts. Blood133, 697–709 (2019).

26. Mestermann, K. et al. The tyrosine kinase inhibitor dasatinib acts as a pharmacologic on/off switch for CAR T cells. Sci. Transl. Med.11, eaau5907 (2019).

27. Liu, P. et al. Nivolumab treatment for relapsed / refractory Epstein-Barr virus-associated hemophagocytic lymphohistocytosis in adults. Blood135, 826–833 (2020).

28. Goldbach-Mansky, R. et al. Comparison of Tripterygium wilfordii Hook F versus sulfasalazine in the treatment of rheumatoid arthritis: A randomized trial. Ann. Intern. Med.151, 229–240 (2009).

29. Wang, S. et al. Tripterygium wilfordii Glycosides Upregulate the New Anti-Inflammatory Cytokine IL-37 through ERK1/2 and p38 MAPK Signal Pathways. Evidence-based Complement. Altern. Med.2017, 1–6 (2017).

30. Singh, N. et al. Monocyte lineage–derived IL-6 does not affect chimeric antigen receptor T-cell function. Cytotherapy19, 867–880 (2017).

31. Zhu, X., Liu, Z., Liu, D., Chen, Z. & Li, L. The effects of triptolide on cell growth and immunologic functions of human dendritic cells. J. Nephrol. Dial. Transpl.10, 217–222 (2001).

32. Hou, W., Liu, B. & Xu, H. Triptolide: Medicinal chemistry, chemical biology and clinical progress. Eur. J. Med. Chem.176, 378–392 (2019).

33. Titov, D. V. et al. XPB, a subunit of TFIIH, is a target of the natural product triptolide. Nat. Chem. Biol.7, 182–188 (2011).

34. Wang, D. et al. Clinical characteristics of 138 hospitalized patients with 2019 novel coronavirus-infected pneumonia in Wuhan, China. JAMA - J. Am. Med. Assoc.323, 1061–1069 (2020).

35. Xu, X. et al. Effective treatment of severe COVID-19 patients with tocilizumab. Proc Natl Acad Sci U S A117, 10970–10975 (2020).

